# Does practice match protocol? A comparison of “triage-to-provider” time among more- vs. less-acute ED patients

**DOI:** 10.1101/2024.02.11.24302630

**Authors:** Temesgen Tsige, Rida Nasir, Daisy Puca, Kevin Charles, Sandhya LoGalbo, Lisa Iyeke, Lindsay Jordan, Melva Morales Sierra, David Silver, Mark Richman

**Author notes:** **Author contributions:** Mark Richman, David Silver, Lisa Iyeke, Lindsay Jordan, Rida Nasir, Kevin Charles, Daisy Puca: Conceived of the project and did the initial manuscript draft and literature search. Temesgen Tsige, Sandhya LoGalbo, Melva Morales Sierra: Created data collection tool, performed data collection and entry, integrated those findings and references into the manuscript, reviewed and edited final manuscript.

## Abstract

**INTRODUCTION:** The Emergency Severity Index (ESI) stratifies Emergency Department (ED) patients for triage, from “most-acute” (level 1) to “least-acute” (level 5). Many EDs have a split-flow model where less-acute (ESI 4 and 5) are seen in a Fast Track, while more-acute (ESI 1, 2, and 3) are seen in the acute care area. As a core principle of Emergency Medicine is to attend to more-acute patients first, deliberately designating an area for less-acute patients to be seen quickly might result in their being seen before more-acute patients. This study aims to determine the percentage of less-acute patients seen by a provider sooner after triage than more-acute patients who arrived within 10 minutes of one another. Additionally, this study compares the Fast Track and acute care areas to see if location affects triage-to-provider time.

**METHODS:** A random convenience sample of 252 ED patients aged ≥18 was taken. Patients were included if their ESI was available for the provider during sign-up. Patients were excluded if they were directly sent to the ED psychiatric area or attended by the author. We collected data on ESI level, timestamps for triage and first provider sign-up, and location to which patient was triaged (Fast Track vs. acute care). Paired patients’ ESI levels, locations, and triage and first provider sign-up times were compared.

**RESULTS:** One hundred twenty-six pairs of patients were included. More-acute patients were seen significantly-faster after triage (∼20 minutes) than less-acute patients in two groups: ESI level 2 vs. 3 and overall high-vs. low-acuity. However, in 34.8% of paired ESI 2 vs. 3 patients, the ESI 3 patient was seen prior to the paired ESI 2 patient, and in 39.4% of overall paired high vs. low acuity patients, the less-acute patient was seen before the more-acute patient. Additionally, patients in the acute care area had significantly-shorter median triage-to-provider times (**∼** 40 minutes) compared to those in the Fast Track area for ESI 2 (acute care) vs ESI 3 (Fast Track) and overall high-acuity (acute care) vs low-acuity (Fast-track). Nonetheless, approximately one-third of ESI 3 patients triaged to Fast Track were seen before ESI 2 patients triaged to the acute care area.

**CONCLUSION:** The split-flow model reduces overall ED length of stay (LOS), improving flow volume, revenue, and patient satisfaction. However, it comes at the expense of the fundamental ethos of Emergency Medicine and potentially subverts the intended triage process. Although most more-acute patients are seen by a provider sooner after triage than less-acute patients, a substantial number are seen later, which could delay urgent medical needs and impact patients’ outcome negatively. Furthermore, acute care area patients are seen sooner post-triage than identical-ESI-level Fast Track patients, suggesting Fast Track might not function as intended. Further examination of patient outcomes is necessary to determine the impact of the ESI triage process and spilt-flow model.

## INTRODUCTION

The Emergency Severity Index (ESI) is a standard means of stratifying Emergency Department (ED) patients for triage, so as to be seen in a particular order (from “most-acute” to “least-acute”)

^1^ESI levels range from 1 (most-acute) to 5 (least-acute). ESI levels 1 and 2 are “emergent” (require immediate assessment and intervention); ESI level 3 is “urgent” (can safely wait a short amount of time); ESI levels 4 and 5 are “non-urgent” (can safely wait a long time)^2^.

The original basis of determining an ESI level was a combination of acuity and the expected number of resources the patient would use in the ED. In practice, an ESI level is assigned based on the gestalt and experience of the person performing triage, supplemented with standardized criteria, such as vital sign abnormalities.^1^

The ethics and imperative of Emergency Medicine require providers to attend to the sickest patients first^3^. It should, therefore, not routinely occur that less-acute patients (ESI level 4 or 5) should be seen sooner after arrival than more-acute patients (ESI levels 1, 2, or 3) who arrive at approximately the same time.

Many EDs have instituted a split flow model^4^ in which potentially less-acute patients (ESI 4 and 5) are seen in a “fast track” (where patients have a lower expected length-of-stay) while more-acute patients (ESI 1, 2, and 3) are seen in a separate area. In this manner, lower-acuity patients can be seen rapidly, which is beneficial to both the patient and the ED. Split flow models have been associated with decreased length-of-stay for low-acuity and for overall ED patients^4^. This can generate substantial revenue from the volume and difference between billing collections and resources expended on low-acuity patients, especially because many EDs charge a facility fee^5^ to each patient, regardless of level of complexity.

The split flow model introduces the possible scenario in which two patients arrive at approximately the same time and have different ESI levels, but the patient determined to potentially have a more-serious condition (ESI 1, 2, or 3) is seen later after arrival than a patient with a likely less-serious condition (ESI 4 or 5) including those seen in a fast track area. This may subvert the ethos/tenet of Emergency Medicine.

This study aimed to determine what percent of less-acute patients have lower triage-to-provider time than more-acute patients and quantify the difference in triage-to-provider time between more-vs. less-acute patients.

## METHODS

The Long Island Jewish Medical Center (LIJMC) is a 583-bed tertiary-care academic hospital, serving a racially- and socio-economically diverse population. The adult ED sees approximately 100,000 patients per year and has an internal area designated for psychiatric patients. The ED has a Fast Track area to which ESI levels 3, 4, and 5, and an acute care treatment area to which all ESI levels, can be sent after triage. Roughly 60% of ED volume is triaged to the Fast Track area, and the remainder to the psychiatric or acute care areas. While the area to which most ESI 1, 2, 4, and 5 patients should be triaged is usually clear (ESI 1 and 2 to acute care, ESI 4 and 5 to Fast Track), this is less clear among the ESI 3 group, which comprises patients with substantial heterogeneity in potential severity (eg, older patients with chest pain; young men with right lower quadrant pain). Consequently, the decision of the area to which each ESI 3 patient should be triaged is more complicated and based on estimated resources needed, professional judgment, and bed availability in various ED areas.

A convenience sample of 126 pairs of patients age ≥18 years was taken between April 24 and December 13, 2023. Any pair of patients whose triage times were within 10 minutes of each other was selected and included as long as the provider sign-up time for each patient was after triage, so the ESI level was available for the provider to see, as that might affect how the provider prioritized which patient to see first. Patients were excluded if they were either directly sent to the psychiatric area of the ED after triage, or if any of the authors was their Attending physician.

We conducted real-time review of the electronic record (EHR) timestamps of patients arriving in the ED, looking for patients with different ESI levels and with triage timestamps within 10 minutes of each other. Patients with ESI 1 or 2 were considered “high-acuity;” those ESI 3, 4, and 5 were considered “low-acuity.” Variables collected were ESI level, ED location to which the patient was triaged (Fast Track vs. acute care area), and timestamps for triage time and time of first provider sign-up; Attending physicians, Resident physicians, physician assistants (PAs), and nurse practitioners (NPs) were considered to be providers. The difference between triage time and provider sign-up time was calculated for each pair of patients. The mean difference in time from triage-to-provider between pairs was determined, as was the percentage of time the less-acute patient of the pair was seen prior to the more-acute patient. The timestamp “triage” (rather than “arrival time” or “room time”) was chosen because triage is the first time during an ED visit a licensed healthcare provider has determined acuity and can expedite care for more-acute patients. We also investigated the influence of triage destinations (Fast Track vs. acute care area) on triage-to-provider time for different ESI levels.

Mann-Whitney test was used to compare median times. Statistical significance was set a priori at p <0.05.

This study was deemed exempt by the Institutional Review Board (IRB #: 23-0169).

## RESULTS

A total of 126 pairs of patients (252 patients overall) with different ESI levels whose triage time was within 10 minutes of each were included. A similar proportion of more-acute and less-acute patients arrived first in the pairing (p = 0.72). (**Table 1**). Forty percent of patients were male and two-thirds were under age 65.

**Table 1.** Percent of times the more-acute patient in the pair arrived before less-acute patient.

| Acuity Level | # of patients that arrived first | % of patients that arrived first |
| --- | --- | --- |
| More acute | 61 | 48.4 |
| Less acute | 65 | 51.6 |

There was a statistically-significant difference (approximately 20 minutes) in triage-to-provider times for paired ESI 2 vs. 3 patients (p = 0.0007) and overall paired high-vs. low-acuity patients (p = 0.004). (**Table 2**) However, in 34.8% of paired ESI 2 vs. 3 patients, the ESI 3 patient was seen prior to the paired ESI 2 patient; and in 39.4% of overall paired high vs. low acuity patients, the less-acute patient was seen before the more-acute patient. While, perhaps owing to small sample sizes of these pairings, no significant difference in median times was found between ESI 2 vs. 4 or 3 vs. 4, in these pairings an even-higher percentage of less-acute patients were seen first: 53.3% and 66.7%, respectively (**Table 3**).

**Table 2.** Triage-to-provider time comparisons for different pairs of ESI levels.

| ESI comparison group | ESI level | Median triage-to-provider time (minutes) (N) | P-value |
| --- | --- | --- | --- |
| 1 vs. 3 | 1 | 19.5 (4) | 0.2 |
|  | 3 | 62 (4) |  |
| 2 vs. 3 | 2 | 35.5 (92) | 0.0007 |
|  | 3 | 60.5 (92) |  |
| 2 vs. 4 | 2 | 39 (15) | 0.69 |
|  | 4 | 27 (15) |  |
| 3 vs. 4 | 3 | 63 (15) | 0.4 |
|  | 4 | 41 (15) |  |
| More-acute vs. less-acute | More-acute | 39.5 (126) | 0.004 |
|  | Less-acute | 55 (126) |  |

**Table 3.** Percent of time the less-acute patient was seen before the more-acute patient in the pair.

| ESI comparison group | Comparison group N | % of pairs in which less-acute patient seen first (N) | Median time difference (minutes) |
| --- | --- | --- | --- |
| 1 vs. 3 | 4 | 0 (0) | 30 |
| 2 vs. 3 | 92 | 34.8 (32) | 13 |
| 2 vs. 4 | 15 | 53.3 (8) | -2 |
| 3 vs. 4 | 15 | 66.7 (10) | -11 |
| More-acute vs. less-acute | 126 | 39.4 (50) | 9 |

More than 74 % of the patients assigned Fast Track were ESI 3 compared to only 22 % in the acute care area; Similarly, in the acute care area 70 % of the patients were assigned ESI 2 compared to 5 % in the Fast Track (**Table 4**).

**Table 4.** ESI level assigned, Fast Track vs. acute care area.

| **ESI Level Assigned** | **% of all patients assigned this ESI level** | **% of acute care ED patients with this ESI level** | **% of Fast Track patients with this ESI level** |
| --- | --- | --- | --- |
| 1 | 1.6 | 3 | 0 |
| 2 | 42.1 | 70 | 5 |
| 3 | 44.4 | 22 | 74 |
| 4 | 11.9 | 5 | 21 |

Median triage-to-provider time among more-acute patients triaged to the acute care ED was approximately 40 minutes less than for less-acute patients triaged to Fast Track (76 vs. 33 minutes, p = 0.0008, for ESI 2 vs. 3; 71.5 vs. 33.5 minutes, p = 0.0004 for overall more-vs. less-acute. (**Table 5**)

**Table 5.** Triage-to-provider time comparisons for different pairs of ESI levels triaged to acute care vs. Fast Track.

| ESI comparison group | ESI level | Median time between triage-to-provider (minutes) (N) | P- value |
| --- | --- | --- | --- |
| 1 (Acute care) vs. 3 (Fast Track) | 1 | 19.5 (2) | 0.33 |
|  | 3 | 80.5 (2) |  |
| 2 (Acute care) vs. 3 (Fast Track) | 2 | 33 (63) | 0.0008 |
|  | 3 | 76 (63) |  |
| 2 (Acute care) vs. 4 (Fast Track) | 2 | 36.5 (8) | 0.64 |
|  | 4 | 42 (8) |  |
| 3 (Acute care) vs. 4 (Fast Track) | 3 | 42 (3) | 1 |
|  | 4 | 29 (3) |  |
| Acute care vs. Fast Track | Acute care | 33.5 (76) | 0.0004 |
|  | Fast Track | 71.5 (76) |  |

*Note:* Location is placed in parentheses; for instance, 2 (acute care) vs. 3 (Fast Track): indicating ESI 2 patients were treated in acute care compared to ESI 3 patients treated in the Fast Track area.

Nonetheless, approximately one-third of ESI 3 patients triaged to Fast Track were seen before ESI 2 patients triaged to the acute care area. (**Table 6**)

**Table 6.** Percent of Fast Track patients with shorter triage-to-provider time compared with acute care area patients for various ESI level groupings.

| **Comparison Group** | **Total comparison** | **% Fast Track patients seen first (N)** | **Median time difference (minutes)** |
| --- | --- | --- | --- |
| ESI 1 (Acute care) vs. ESI 3 (Fast Track) | 2 | 0 (0) | 61 |
| ESI 2 (Acute care) vs. ESI 3 (Fast Track) | 63 | 31.75 (20) | 23 |
| ESI 2 (Acute care) vs. ESI 4 (Fast Track) | 8 | 37.5 (3) | 12 |
| ESI 3 (Acute care) vs. ESI 4 (Fast Track) | 3 | 33.3 (1) | 3 |
| Overall Acute care vs. Fast Track | 76 | 31.6 (24) | 19.5 |

Among ESI level 3 patients generally (not paired, because this study looked at pairs of different ESI levels), patients triaged to the Acute care area median triage-to-provider time was 30 minutes sooner than those triaged to the Fast Track (p = 0.008). (**Table 7**)

**Table 7.** ESI 3 Triage-to-provider Times for Fast Track vs. Acute care area.

| **Location** | **Median triage-to-provider time (minutes)** | **P-value** |
| --- | --- | --- |
| Acute care area | 46 | 0.008 |
| Fast Track | 76 |  |

## DISCUSSION

The split flow model of Emergency Department care has been associated with decreased overall length of stay (LOS), both among patients triaged to the low-acuity area and those triaged to the higher-acuity areas.^4^ This is done to increase overall ED flow volume, revenue, and patient satisfaction. However, this study reveals unintended consequences of the split flow model. Namely, it comes at the expense of the fundamental ethos/tenet of Emergency Medicine and subverts the intended triage process in such a way that a substantial number (∼40% in this study) of patients deemed by the ESI triage level to be of lower-acuity are seen before patients triaged as higher-acuity. This study unveils the tension between ED’s financial and patient satisfaction goals to see less-acute patients quickly vs. the ethical responsibility to see more-acute patients quickly.

Not surprisingly, the study found more-acute patients were seen significantly faster than less-acute patients in two different comparison groups: ESI 2 vs. 3 (the most-common comparison pair) and overall high-vs. low-acuity groups. However, even within these groups, a large percentage of less-acute patients (34.8% and 39.4% respectively) were seen before more-acute patients. Similar findings were also observed in other comparison groups, but not at statistically-significant levels. While the overall trend favors more-acute patients being seen sooner after triage, the high percentage of less-acute patients seen before more-acute patients is potentially harmful, as it could delay care for those with urgent medical needs and negatively impact their outcomes.

There was an interesting distribution of ESI levels assigned in the Fast Track and acute care ED areas. More than 74 % of patients assigned to the Fast Track were ESI 3, compared to only 22 % in the acute care area. Similarly, 70 % of patients in the acute care area were assigned ESI 2, compared to only 5 % in the Fast Track. This suggests the Fast Track might not be acting as a “fast track” for such patients.

Additionally, patients in the acute care area were seen significantly sooner post-triage than those in the Fast Track area in two comparison groups: ESI 3 (Fast Track) vs. ESI 2 (acute care ED) and overall less-acute (Fast Track) vs. more-acute (acute care area). This finding raises questions about how effective the split-flow model, particularly the Fast Track area, is in achieving its intended goal of allowing less-acute (less sick) patients to be seen by a provider faster. Even among ESI level 3 patients only, patients triaged to Fast Track were seen 28 minutes later than those triaged to the acute care ED (p = 0.008). This finding further strengthens the evidence that the Fast Track isn’t functioning as a “fast track.” The heterogeneous/diverse clinical profiles among ESI 3 patients may contribute to their longer wait times to be seen in the Fast Track area^6^.

However, the observed longer wait times for patients in the Fast Track setting suggest a potential need for a better triage method. Research suggests that certain patient groups, including those from low-income backgrounds or ethnic minorities, may be disproportionately mistriaged in the ED^7^. There is a need for additional tools or methods that consider patients holistically and integrate factors beyond symptoms and vital signs. Improving triage protocols might increase placement of patients to the appropriate setting (eg, Fast Track or acute care area), while improving resource allocation and promoting equitable access to care. Such a triage process has been implemented, leveraging artificial intelligence to produce risk-driven triage acuity suggestions embedded in the EHR.^8,9,10^ Results indicate a decrease in the percent of patients triaged as level 3 and an increase in levels 4 and 5, without increasing risk or length of stay for the low-acuity groups.

While the ED should try, as often as possible, to see more-acute patients sooner from triage than less-acute patients, it would not be reasonable to suggest every more-acute patient be seen sooner. This is because, while less-sick patients are waiting to be seen, more-sick patients will likely arrive, thereby relegating less-sick patients to the “back of the line” over and over. Several options might relieve this conflict between the desire to see lower-acuity patients quickly and the imperative to see higher-acuity patients quickly:

1. Institutions should consider redistributing their workforce such that providers are active in the triage process (ie, greatly minimize the time from “arrival to provider”).
2. EDs can eliminate the split flow model and incorporate both less-acute and more-acute patients in the same areas of the ED, so providers can see a mix of both levels of patients.
3. Organizations can determine less-sick patients should be expected to wait a reasonable period of time (perhaps two hours) before being seen. Once they reach the two-hour limit, a provider could be notified to see them expeditiously, unless they are actively occupied with a more-sick patient (ie, not just waiting for the next more-sick patient to be brought to their treatment area).
4. Finally, owing to the Emergency Medical Treatment & Labor Act (EMTALA), EDs are obliged to provide patients a medical screening examination (MSE) to determine level of sickness^11^; while the practitioner conducting the MSE need not be a licensed independent provider (but, rather, “medical personnel … qualified to perform an MSE that is appropriate to the individual’s presenting signs and symptoms,”^12^ in practice, it is almost always such a provider who performs the MSE, which often becomes a full assessment and treatment visit, even for minor complaints. In New York, such providers can rapidly discharge patients, either to home or to rapid follow-up at a facility for lower-acuity visits (e.g., urgent care centers), including one owned and operated by the same corporation that owns/operates the ED.^9^

## LIMITATIONS

This study has several limitations. First, it was a single-center study; results may differ at other institutions, particularly because LIJ ED’s triage patterns may not reflect those in other EDs. A very high percentage (74%) of LIJ’s Fast Track patients are ESI 3, and up to 60% of the ED volume comes through Fast Track, for a total of ∼45,000 Fast Track ESI 3 patients/year. Other EDs may reserve their Fast Track areas primarily for ESI 4 and 5 patients. Second, we used the timestamp of provider sign-up to indicate when a provider first saw the patient. However, sometimes a provider might sign up for a patient shortly after they are triaged to that provider’s area, without seeing the patient until the patient arrives in the provider’s area. In such circumstances, signing up for the patient indicates not the “time seen,” but, rather, the time the provider took responsibility for the patient. Finally, this study did not look at outcomes. The next step in our research will be to examine outcomes (total ED LOS, disposition (eg, admit vs. discharge) and 30-day return visits) of patients in our study.

## CONCLUSION

The ESI triage system, as currently utilized, largely succeeds in ensuring more-acute patients are seen sooner after triage than less-acute patients. However, a substantial percent of less-acute patients are still seen earlier post-triage than more-acute patients. This phenomenon is worthy of additional studies at different institutions and following-up patients to determine whether it is associated with adverse outcomes.

## Patient’s Consent

N/A

## Trial/Systematic Review Registry

N/A

## Data Availability

All data produced in the present study are available upon reasonable request to the authors.

## Acknowledgements

None.

## ETHICAL APPROVAL

As a quality improvement project, this study was deemed not to meet the definition of research by the Institutional Review Board in its Human Subjects Determination Request evaluation.

## CONFLICT OF INTERESTS

## FUNDING SUPPORT

## AVAILABILITY OF DATA AND MATERIALS

## Notes

### Competing Interest Statement

The authors have declared no competing interest.

### Funding Statement

This study did not receive any funding.

### Author Declarations

IRB of Northwell Health Long Island Jewish Medical Center gave ethical approval for this work.

## REFERENCES

1 Gilboy N, Tanabe P, Travers D, Rosenau AM: Emergency Severity Index (ESI): A triage tool for emergency department care (Version 4). Schaumburg, IL: Emergency Nurses Association. 2020, https://media.emscimprovement.center/documents/esi-implementation-handbook-2020_1592279717486_edvzge5cc.pdf

2 Reiter M, Scaletta T: On Your Mark, Get Set, Triage!. Emergency Physicians Monthly | Independent news and analysis in emergency medicine. 2008, https://epmonthly.com/article/on-your-mark-get-set-triage/

3 Yancey CC, O’Rourke MC: Emergency Department Triage. In StatPearls. StatPearls Publishing. 2023, http://www.ncbi.nlm.nih.gov/books/NBK557583/

4 Pierce BA, Gormley D: Are Split Flow and Provider in Triage Models in the Emergency Department Effective in Reducing Discharge Length of Stay? Journal of emergency nursing, 2016, 42(6), 487–491. 10.1016/j.jen.2016.01.005

5 Claxton G, Cox C, Rae M, Schwartz H: How do facility fees contribute to rising emergency department costs? Peterson-KFF Health System Tracker. 2023, https://www.healthsystemtracker.org/brief/how-do-facility-fees-contribute-to-rising-emergency-department-costs/

6 Hinson J S, Levin S: Data-Driven Approach Yields New Approach for Emergency Department Triage In ACEP Now. 2023, https://www.acepnow.com/article/data-driven-approach-yields-new-approach-for-emergency-department-triage/2/

7. Sax DR, Warton EM, Mark DG, et al. Evaluation of the Emergency Severity Index in US Emergency Departments for the Rate of Mistriage. JAMA Network Open. 10.1001/jamanetworkopen.2023.3404

8. Dugas AF, Kirsch TD, Toerper M, et al. An Electronic Emergency Triage System to Improve Patient Distribution by Critical Outcomes. Journal of Emergency Medicine. 2016;50(6):910–918.

9 Success Stories From the AHRQ Digital Healthcare Research Program – HopScore: An Electronic Outcomes-Based Emergency Triage System. Published online January 23, 2020. Accessed January 23, 2020.

10. Levin S, Toerper M, Hinson J, et al. Machine-learning-based electronic triage: a prospective evaluation. Annals of Emergency Medicine. 2018;72(4).

11 Emergency Medical Treatment & Labor Act (EMTALA) | CMS. https://www.cms.gov/medicare/regulations-guidance/legislation/emergency-medical-treatment-labor-act

12 The New York State Department of Health. 2020, https://www.health.ny.gov/professionals/hospital_administrator/letters/2020/docs/dal_20-07_emtala.pdf10. Levin S, Toerper M, Hamrock E, et al. Machine-learning-based electronic triage more accurately differentiates patients with respect to clinical outcomes compared with the emergency severity index. Annals of Emergency Medicine. 2018;71(5):565–574.

